# The Interplay of Spirituality and Self-Regulation in Youths: A Real-Time Examination of Mental Health Dynamics (SPIRIT)

**DOI:** 10.64898/2026.03.27.26349490

**Authors:** Sébastien Urben, Chloé Von Niederhäusern, Setareh Ranjbar, Kerstin Jessica Plessen, Jennifer Glaus

## Abstract

**Background:** Adolescence and young adulthood represent critical developmental stages during which mental disorders often emerge, with the potential to impede perceived quality of life. Spirituality (i.e., the search for the sacred) and self-regulation (i.e., intrinsic processes regulating emotions, thoughts, and behaviors) are recognized as protective factors for mental health. However, their dynamic interplay remains largely unexplored, particularly in real-life and in real-time among youths. This study, developed with the help of young partners, addresses this gap by investigating the longitudinal associations between spirituality, self-regulation, and mental health using an ecological momentary assessment (EMA) approach.

**Methods and analysis:** We plan to recruit 120 adolescents and young adults (aged 16–20, expected attrition rate of 20%) from the community to complete a qualitative semi-structured interview assessing their beliefs, spiritual or religious activities, role models, and “meaning in life”. In addition, participants will take part in a multi-wave intensive longitudinal study. Trait-level assessments will be conducted at two time points, three months apart, to capture between-person differences. Additionally, to assess within-person dynamics, participants will complete EMA surveys four times daily over 10 consecutive days in two waves, also three months apart. Measures will include facets of spirituality (e.g., beliefs, meaning, collective consciousness), self-regulation (e.g., self-control, emotional regulation, impulsivity), as well as mental health indicators (emotional and behavioral symptoms) and quality of life. Qualitative data will be analyzed through a thematic analysis method, whereas quantitative associations will be assessed using Linear Mixed Models (LMM) and network analyses.

**Ethics and dissemination:** Ethical approval has been obtained, and data collection begun in May 2025. Findings will be disseminated through open access peer-reviewed journals, conferences on adolescent mental health, and shared with practitioners, educators, and youth organizations. Results will also be made accessible to the general public. This study aims to inform personalized preventive and therapeutic interventions by elucidating real-time mechanisms linking spirituality, self-regulation, and mental health in youths.

**Strengths and limitations of this study:**

- This study design combines qualitative and quantitative approaches.
- We use real-time, multi-wave ecological momentary assessment (EMA) to examine spirituality, self-regulation, and mental health in youths.
- We capture both within- and between-person variations, providing a fine-grained understanding of the temporality and dynamics of these processes.
- We integrate multiple dimensions of spirituality, self-regulation, and mental health rather than focusing on single constructs.
- Real-life and real-time assessments reduce recall bias and increase relevance to daily experiences.
- We invited a panel of youths from the general population to provide feedback during the development phase of the project.
- The results may not be generalizable to younger adolescents or older adults as the age range is restricted to 16–20 years.
- Reliance on self-reported data may introduce bias (e.g., social desirability, subjective interpretation).

## 1. Introduction

### 1.1 Integrative approach of mental health

Adolescence or emerging adulthood is a critical and vulnerable period when the most serious mental disorders typically emerge^1^. This is a period of transition during which problems of mental health may arise, potentially resulting in a perceived decline in overall quality of life^2^.

In this context, an integrative and holistic approach to the mental health of youths allows to focus on care and self-care putting forward the psychological, physical, and spiritual needs of the individual^3^. In this perspective, the bio-psycho-social approach, enriched by the contribution of spirituality, is essential in understanding both somatic^4^ and mental health^5^. This is in line with a recent Swiss population-based survey^6^, showing that 79% of the respondents would like their health care professional to use an integrative and holistic approach to meet their health care needs. Similarly, in 2019, a Swiss national survey showed that 40% of people believe in a single God and 51% believe that a higher force guides our destiny^7^. Moreover, recent data from the Swiss Federal Statistical Office in 2024 indicate that 56.1% of respondents consider religion and spirituality important during challenging life situations, and 49.1% of individuals aged 15–24 engage in prayer practices^8^.

### 1.2 Spirituality

We defined spirituality as “*the search for the sacred*”^9^, with the sacred relating to “*aspects of life that are perceived as manifestations of the divine or imbued with divine-like qualities, such as transcendence, immanence, boundlessness and ultimacy*”^9^. Thus, spirituality refers to a process allowing people to discover what is of ultimate importance to them in life. It has personal, interpersonal, and transpersonal dimensions consisting, mainly, of four components^10^: (a) a belief in a higher power or universal intelligence; (b) an inner search for meaning and purpose (i.e., self-discovery process); (c) authentic connections to others based on deep respect and reverence for life; and (d) an eco-awareness which represents a connection to nature, respect for the environment, and the belief that the Earth is sacred.

Spirituality includes, but is not limited to, religion or religiosity. Indeed, religiosity refers to narrower concepts that consider practices, beliefs, and symbols organized into a system of faith^11^. It is important to note that spirituality and religiosity have different impacts on individuals. Religiosity appears to be beneficial in the adoption of better lifestyle habits, including diet, tobacco, and alcohol consumption. Thus, the effects of religiosity on health may be attributed to the behavioral regulation of health habits^12^. In contrast, spirituality has been linked to an increased sense of meaning, purpose, resilience, satisfaction, happiness, and well-being^13–15^, and provides meaning and support in times of stress^16–18^. In particular, spirituality has been associated to biomarkers of health and somatic disease progression, such as blood pressure, cardiac reactivity, and immune system markers^12^. Therefore, systematic reviews and meta-analyses demonstrated that spirituality represents a protective factor for mental health^19–24^ especially in youths^16,25^.

Building on this multidimensional view of spirituality, it has been proposed^26^ that religious traditions and spirituality provide five complementary psychological resources that support coping and meaning-making: (1) emotional–affective: experiences of transcendence and emotional regulation; (2) psychosocial: social support and belonging; (3) cognitive: coherent worldviews for interpreting life events; (4) ethical: moral values and behavioral guidelines for self-regulation; and (5) identity: roles and exemplars that help individuals situate themselves in the world. Together, these resources form a flexible constellation that individuals can draw upon according to their needs, offering a comprehensive framework for understanding how spirituality promotes well-being.

Evidence suggests that the impact of spirituality on health is primarily mediated through physiological processes sustaining emotion regulation, or more largely self-regulation^12^. Self-regulation is an umbrella term for intrinsic mechanisms that govern emotions, thoughts, and behaviors, often aligning immediate actions with long-term goals^27^, and it constitutes a critical protective factor for mental health. Among Indian and Emirati adolescents, self-control (a key aspect of self-regulation) has been shown to moderate the relationship between spirituality and internalized symptoms, as well as life satisfaction (or quality of life)^28^.

### 1.3 Qualitative approach

Qualitative research on youth spirituality has mainly evolved along two lines: clinical/health-related settings and cultural environments where religion and spirituality are particularly salient. In health-related studies, spirituality is often conceptualized as a coping resource, particularly through hope, as illustrated by qualitative work with adolescents and young adults living with cancer^29^ and by studies linking spiritual health to hope among adolescents^30^. More exploratory studies have also examined how adolescents define spirituality and what shapes these meanings (e.g., purpose, relationships, values, everyday practices), nevertheless, all existing studies have been conducted in contexts where religion or spirituality holds a specific culturally prominent role compared to European countries^30–32^.

In Switzerland, qualitative studies have largely focused on specific groups. For example, research with Muslim and Buddhist youths highlights religious individualization and efforts toward social recognition and civic participation^33^. In addition, qualitative work with Swiss secular individuals (i.e., individuals without religious affiliation) further shows that spirituality can be articulated beyond religious affiliation as a multidimensional construct^34^. However, to the best of our knowledge, the literature lacks qualitative evidence on how spirituality is experienced and interpreted in the everyday lives of Swiss adolescents from the general population.

### 1.4 Ecological momentary assessment

Moreover, previous knowledge of the association between spirituality, self-regulation, and mental health has been mainly developed through quantitative assessments with poor ecological validity, cross-sectional design, and lack of attention to within-person variations. Ecological momentary assessment (EMA) refers to frequent and repeated measurements in real-life and in real-time^35^ allowing to examine the dynamic interrelationships between spirituality, self-regulation and mental health. In this perspective, EMA has been used to study spirituality^36–44^, mainly among American participants, and in adult community samples^37–42,44^. For instance, the SoulPulse project^37–41^ had a major impact on the study of spirituality in real-time by examining daily spiritual experiences among adults. Indeed, this project observed that daily spirituality served as a buffer against daily stressors^39^, moderated the relationship between stressful life events and well-being^38^, increased well-being both at the state and trait level^38^, and promoted feelings of love and care for others in stressful situations^37^. Awareness of a higher being in everyday life was also observed^40^ to precede feelings of meaningfulness^41^. The only non-American EMA study on spirituality was conducted in a Dutch clinical sample of adults and found that affective representations of God and spiritual experiences varied significantly within the day^43^. Only one EMA study^36^ in USA had sampled adolescents and found that after Ramadan, adolescents felt more connected to Allah in their daily lives and had a lasting effect toward more self-control and patience^36^. In this line, daily diary studies involving adolescents with cystic fibrosis found that engagement in spirituality resulted in greater treatment adherence^45^, and that imbuing their own body with divine significance (i.e., body sanctification) led to better sleep hygiene and other health-protective behaviors^46^.

### 1.5 Research gaps

The literature on qualitative studies of youth spirituality remains scarce and has predominantly focused on highly religious or clinical populations, such as adolescents with chronic illnesses^47^, or those living in strongly religious cultural contexts^30–32^. While these studies provide important insights into how young people understand spirituality, they do not address the everyday spiritual experiences of youths from the general population in secular European settings. As a result, the field still lacks systematic qualitative accounts of how young people experience, articulate, and interpret spirituality in their daily lives. This gap is especially salient in Switzerland, where a culturally diverse religious and spiritual environment may shape both the content of spirituality and its associations with well-being, calling for qualitative inquiry that is sensitive to contextual and multidimensional aspects of lived experience^48,49^.

In addition, the existing literature does not, to the best of our knowledge, combine such qualitative perspectives with intensive quantitative assessments, this design is therefore well suited to clarify how spirituality relates to self-regulatory processes and mental health in the natural environments of Swiss youths. In this perspective, the use of multiple waves of EMA contributes to the accuracy and diversity of daily observations by capturing different and distant slices of life^35^. However, all previous studies had a single-wave EMA approach^37–44^ except the Ramadan study, which used a three-wave EMA^36^. Therefore, intensive longitudinal studies are needed to provide additional evidence and a greater degree of proof of spirituality as a protective factor in real-life and real-time among youths from the general population. It is important to note that these concepts encompass various components that have not been thoroughly explored until now. All of the mentioned EMA studies show a predominant focus on adults and on the American population. Importantly, none of these studies evaluated spirituality, self-regulation, and mental health together and across their different components. Therefore, the understanding of the real-time, dynamic associations between the various facets of spirituality, self-regulation, and mental health as well as quality of life at the within-person level and in a real-life context is still largely unexplored in European adolescent and young adult populations. Furthermore, no studies exist in Switzerland, yet, despite the fact that the majority of the Swiss population^6^ would like their health care professionals to take this aspect into account.

### 1.6 The current research project

Our overall objective is to gain a deeper understanding of how the components of spirituality and self-regulation impact adolescent and young adult mental health in real-life and in real-time. More specifically, we expect to gain important insights at both the between- and within-person levels, into the specific roles played by distinct components of spirituality and self-regulation in their associations with the mental health and quality of life of youths.

In particular, the aim of this study is to gain a deeper understanding of youths mental health by exploring the qualitative, as well as the quantitative dynamic and longitudinal associations of different facets of spirituality, encompassing the emotional–affective, psychosocial, cognitive, ethical, and identity related dimensions, in conjunction with the components of self-regulation (e.g., self-control, emotional regulation, impulsivity, sense of self-efficacy), mental health (e.g., psychopathological symptoms) and quality of life.

## 2. Methods and analysis

This study combines qualitative alongside quantitative observations trough a two-wave intensive-longitudinal measures. We plan to recruit adolescents and young adults in the community of the Lausanne region in the French speaking part of Switzerland. The study is currently in the data collection phase. It was approved by the Ethics committee of the Vaud Canton, Switzerland (#2025-00088) and is in accordance with the Helsinki Declaration.

### 2.1 Participants

We will recruit 120 participants aged between 16 and 20 years. The inclusion and exclusion criteria are described in Table 1.

**Table 1.** Inclusion / exclusion criteria for children and their legal guardian.

| *Criteria* |  |
| --- | --- |
| *Inclusion* | • Speaking fluent French<br>• Having a smartphone with access to internet data and a Swiss phone number<br>• Being aged between 16 and 20 years<br>• Written informed assent or written informed consent<br>• Known significant difficulties in intellectual, conceptual, practical, or social adjustment that indicate a potential mild or more severe disorders of intellectual development (International Classification of Diseases ICD, 11<sup>th</sup> revision [ICD11], 06A00)<sup>76,77</sup> |
| *Exclusion* | • Known significant difficulties in reading that indicate a potential developmental learning disorder with impairment in reading (ICD11, 6A03.0)<br>• Known major life-crisis (e.g., active suicidal behaviors or thoughts, acute or florid psychosis)<br>• Known significant difficulties in intellectual, conceptual, practical, or social adjustment that indicate a potential mild or more severe disorders of intellectual development (International Classification of Diseases ICD, 11<sup>th</sup> revision [ICD11], 06A00)<sup>76,77</sup> |

### 2.2 Procedure

The study will be conducted in four main steps: (1) qualitative interview; (2) online baseline measures; (3) a 10-day period of EMA; (4) a 3-month follow-up assessment alongside a second 10-day period of EMA.

The protocol procedure is summarized in Figure 1.

**Figure 1.**
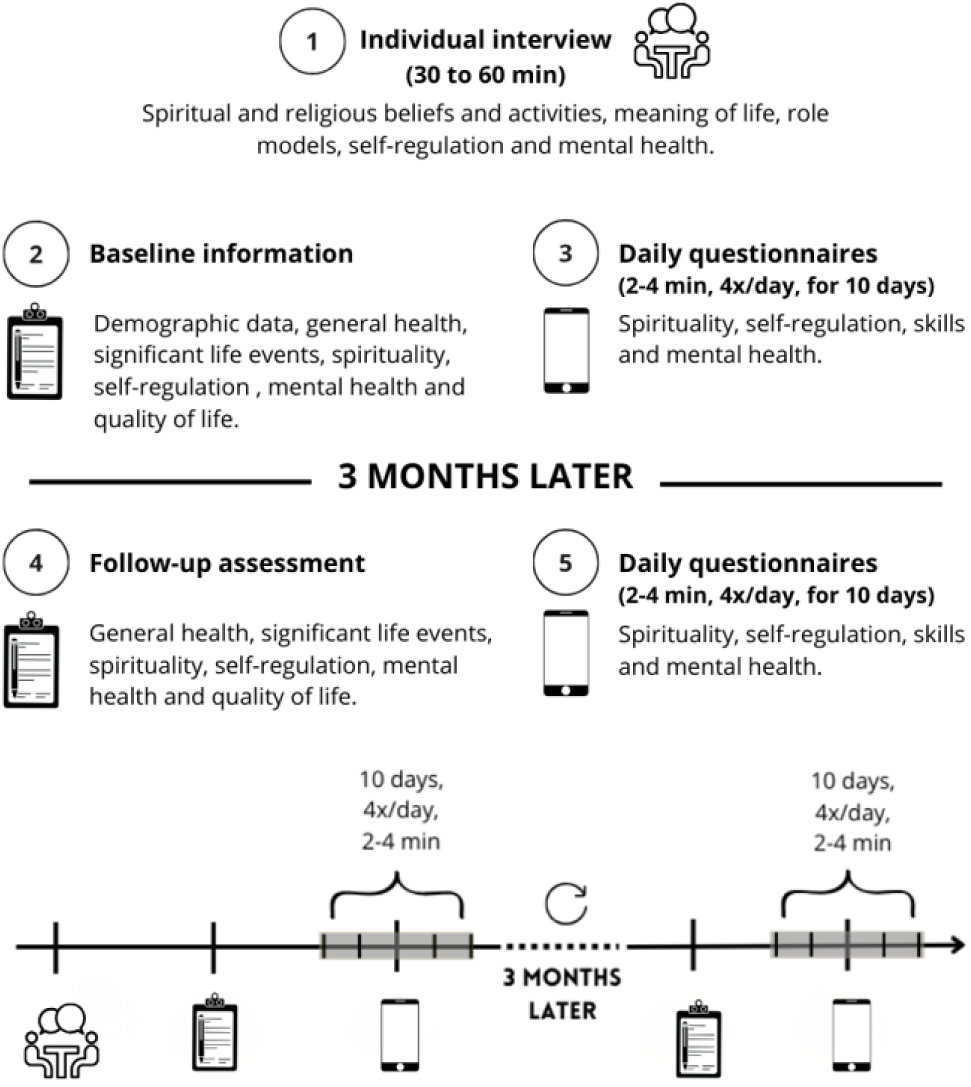
Study procedure.

### 2.3 Measures

#### 2.3.1 Qualitative assessment

We will conduct a qualitative semi-structured interview (either face-to-face or via Webex^©^). We will ask participants to talk about their beliefs, spiritual or religious activities, role models, and meaning in life. This interview will last for 30 minutes to an hour. The interview will be recorded for analyses purpose. The verbatim will be transcribed and double checked for quality. The interviewer will be a psychologist or a master student in psychology. The main themes that will be addressed during the interview are related to spirituality and resources the participants use to cope with their everyday life and to support their daily well-being.

#### 2.3.2 Quantitative assessments

##### Baseline measures

Participants will be asked to complete an online battery of 12 questionnaires about socio-demographics and general health status, life events, spirituality, self-regulation and mental health. All measures will be administered online in French. All questionnaires possess good psychometric properties. The questionnaires not available in French language have been translated from English into French by native speakers well-experienced with clinical diagnostic instruments and back translated by an English native speaker. Individual response timing will automatically be registered to detect outliers in response behavior. The collected EMA data will be stored in a secure web application hosted on the Lausanne University Hospital secured servers (REDCap^®^ ^50,51^) directly upon the completion of each baseline questionnaire or EMA survey, which will ensure the protection of the privacy and the data.

Sociodemographic and general health status variables include questions on age, sex, parental education and profession, cultural background and family constellation as well as religious confession and history, belief in a single God or higher force that guides our destiny. Moreover, information on possible treatments, medication, and somatic issues will be collected.

The Adverse Childhood Experience (ACE) section of the Pediatrics ACE and related life events screener (PEARLS)^52,53^ is a screening tool that assesses *abuse, neglect and family dysfunction*.

Concerning spirituality, we will use three self-report measures: (a) the Spirituality Scale (SS)^10^, which consists of 4 dimensions^54,55^: *spiritual beliefs, self-discovery, self-awareness and collective consciousness, respect for others and the environment*; (b) the Attitudes Related to Spirituality Scale (ARES)^56^, which focus on *non-tautological aspects*. (c) the Meaning in Life Questionnaire (MLQ)^57^ examines two dimension of meaning in life: the *presence of meaning* (how full participants feel their lives are of meaning) and the *search for meaning* (how engaged and motivated participants are in efforts to find meaning or better understand the meaning in their lives).

Self-regulation will include: (a) the Cognitive Emotion Regulation Scale (CERQ)^58^ identifying *adaptive and non-adaptive emotion regulation strategies*; (b) *trait self-control skills* will be assessed with the French version of the Brief Self-Control Scale (BSCS)^59^, (c) *impulsivity* will be assessed using the short French version of the UPPS-P Impulsive Behavior Scale (UPPS)^60,61^ questionnaire that evaluate 5 impulsivity facets, including negative urgency, positive urgency, lack of premeditation, lack of perseverance, and sensation seeking; (d) *self-efficacy* will be measured using the French adaptation of the General Self-Efficacy Scale (GSES)^62^, evaluating self-beliefs to cope with difficult demands in life.

Concerning *mental health*: the French version of the Achenbach System of Empirically Based Assessment, Child Behavior Checklist (CBCL) - Youth Self-Report (YSR)^63,64^ will be used to assess behavioral and emotional problems, including eight categories: anxious/depressed, withdrawn/depressed, somatic complaints, social problems, thought problems, attention problems, rule-breaking behavior, and aggressive behavior. The Positive Mental Health (PMH) Scale^65^ will be used to assess mental well-being.

The KidScreen-27^66–68^ will be used to assess five dimensions of *quality of life*, including physical and psychological well-being, relationships with parents and autonomy, social support and peers, and school environment.

##### Ecological momentary Assessment (EMA)

Participants will receive a short training on how to answer questions from the EMA questionnaire. They will then answer the questions in the programmed questionnaire from their personal smartphone that prompts the participants at given time intervals. Participants will receive an SMS with a website link (within the REDCap^®^ environment^50,51^) where they will be able to answer the questions. Options to postpone the alarm/filling out the questions are available if participants are in a situation in which it is not possible to respond promptly. However, after a period of one hour it is not possible for the participants to respond to the survey anymore (to avoid retrospective assessments). Previous studies^69^ and our own experiences (mSanté study^70^ and ECOSREXT study^71^) have shown that the feasibility of EMA assessment is high (compliance above 70%). The average time to fill out the questionnaire each time is short (2-5 minutes).

More specifically, participants will be asked to answer the EMA surveys 4 times a day for 10 consecutive days at two different periods which will be 3 months apart: a morning survey (29-37 items before school/work), a noon and an afternoon surveys (27-33 items after school/work), and an evening survey (38-45 items before bedtime). The surveys cover the domains of momentary context, emotional states, substance use, daily spirituality, self-control and impulsivity. The evening survey further includes questions on daily life events, quality of life and overall health. The surveys also include some features that we hope will make filling out the questionnaires more enjoyable, such as icons for each question, progress bars, colors, and an avatar chosen by the participants with messages.

##### Follow-up assessment

Before the completion of the second EMA period, which is 3 months after the first EMA period, participants will receive the invitation to a survey. This consists of a follow-up of all the baseline questionnaires, except for the socio-demographics, the general health status and the PEARLS-ACE. We will replace the PEARLS-ACE by a non-validated questionnaire composed of 3 questions asking whether participants have experienced a major life event during the last three months (from the baseline assessment to the follow-up assessment). If yes, what and when.

All the instruments that will be used throughout the study are summarized in Table 2.

**Table 2.**
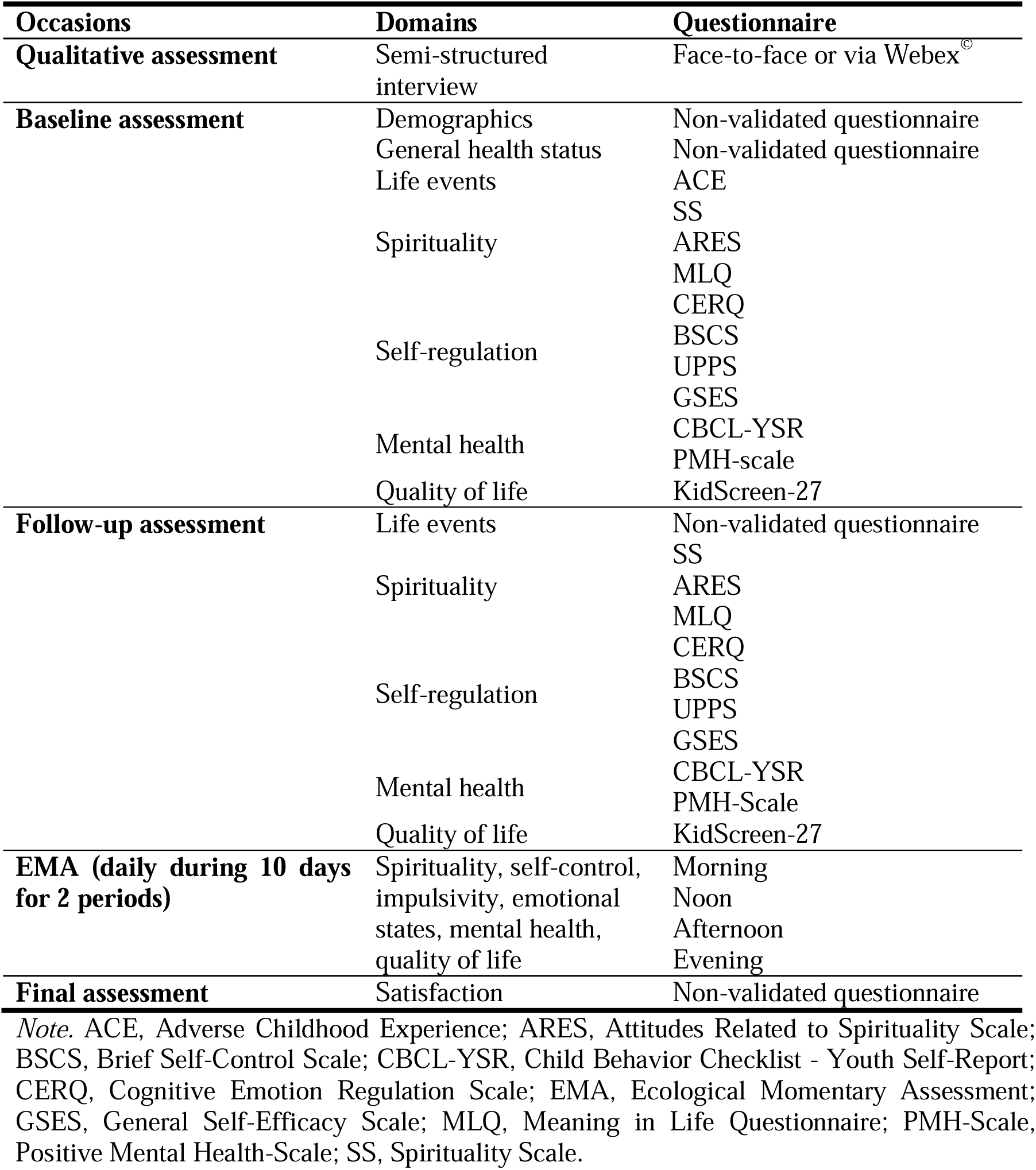
Assessments.

### 2.4 Statistical analysis plan

#### 2.4.1 Data analysis

##### Qualitative approach

Transcription of verbal data will be analyzed using thematic analysis method for identifying and reporting themes within data^72^. Thematic analyses will be carried out by at least 2 experts and their results compared. Any differences between experts will be discussed to achieve a consensus.

##### Quantitative data

Linear Mixed Models (LMM) will be used to study the lagged and cross-lagged associations between daily spirituality, daily self-control, and momentary emotions on well-being, including baseline measures (e.g., self-regulation and spirituality) at the within-and between-person levels, using the daily data collected through EMA for both waves. The LMM are the processes of fitting the following two-level hierarchical regression for each wave:

Level 1 (within-person level): Daily well-being_itw_ = *γ_0iw_* + *γ_1iw_* Daily spirituality_i(t-1)w_ + *γ_2iw_* Daily self-control_i(t-1)w_ *+ γ_3iw_* Momentary emotions_i(t-1)w_ *+ γ*_4iw_ Daily well-being_i(t-1)w_ + *E_itw_*
Level 2 (between-person level): *γ*_0iw_ = *β*_00w_ + *β*_01w_Quality of life_iw_ + *β*_02w_Mental health_iw_ + *β*_03_Spirituality_iw_ + *β*_04_Self-regulation_iw_ + *R*_0iw_

The index (it) indicates the observations for participant (i) and time (t), and (i(t-1)) refers to the observation for the same participant, same wave, at the previous assessment. In addition, the subscript (w) indicates the way and will allow the within-person and between-person effects to vary from one wave to the next, better accounting for the two different time windows in participants’ lives. Level-1 (within-person) and Level-2 (between-person) coefficients will be estimated simultaneously using a multilevel modeling framework (i.e., Linear Mixed Models – LMM). The estimation will rely on the Restricted Maximum Likelihood (REML) method, incorporating both fixed effects and random effects to account for individual variability. This approach allows for modeling the nested structure of the data and accurately representing within-person variation across time.

To exploit the full potential of this intensive longitudinal database, dynamic network analysis will be implemented by estimating a multilevel vector autoregression model on the EMA data^63^ to shed light on the evolution and interplay of well-being, spirituality, self-control, and momentary emotions (measured 4x/day for 10 days). This will result in three different network architectures: (a) the temporal within-person network (lagged model, from one measure to the next one, but also across waves); (b) the contemporaneous within-person network (from a post-hoc analysis); (c) the between-person network structure.

#### 2.4.2 Sample size rationale

The power analysis is conducted by running 1000 simulations based on the LMM design with the following settings: (1) the interclass correlation coefficient (ICC) was fixed at 0.75; (2) small effect sizes based on Cohen’s guidelines for standardized beta are considered at 0.21 for daily spirituality assessments. This analysis shows sufficient power (over 80%) to detect the effect of spirituality on daily well-being, with 10 days (one wave of 10 days) of observation across 100 patients at the 5% significance level. With 20% rate of attrition recruitment of 120 participants are planned. The analyses will be conducted using R^73^ and SAS^74^ softwares.

## 3. Patient and public involvement (PPI)

We invited a panel of youths from the general population to provide feedback during the development phase of the project. They reviewed the qualitative semi-structured interview, the study procedure for the quantitative part of the study (i.e., length of EMA period and length of EMA questions), and the gamification elements planned for the EMA component (e.g., fun facts, choice of avatar) aimed at enhancing participant engagement and adherence. Moreover, their feedback was used to refine the spirituality-related interview questions, ensuring they were clear, concise, and easy for participants to understand. We also asked the youth panel for input on how to report individual results back to participants. Finally, we intend to involve PPI partners in disseminating the study findings to the general public.

## 4. Ethics and dissemination

### 4.1 Ethics

This study adheres to the approved protocol, the Declaration of Helsinki, Good Clinical Practice guidelines, the Human Research Act (HRA), the Human Research Ordinance (HRO), and all applicable local regulations. Ethical approval was obtained from the competent local ethics committee (CER-VD, #2025-00088).

### 4.2 Dissemination

#### 4.2.1 Publication

The findings from this study will be published in open access peer-reviewed journals and presented at leading conferences on child and adolescent mental health, as well as meetings dedicated to ambulatory assessment. Results will also be shared with practitioners through the institute’s strong connections with clinics, educational institutions, and third-sector organizations. Furthermore, we aim to make the outcomes accessible to the public via online and print media public events, and social media platforms.

#### 4.2.2 Data sharing policy

Journals and funding agencies can request the sharing of data from publications on a dedicated website (e.g., Zenodo^©^). This data sharing approach allows the validation of published results (reproducibility), the aggregation of data from different research projects, and more generally the use of the data by other researchers. In the case of such sharing, study participant data will always be coded in such a way that it cannot be traced back to the participant’s identity. The main analyses will be registered on Open Science Framework (OSF).

## 5. Project’s impact

This study has the potential to make a significant impact in several areas. By adopting an innovative and original integrative approach, it will allow a fine-grained observation of the complex longitudinal dynamic associations between different components of spirituality, self-regulation and mental health. The findings of the study may help implement positive attitudes in adolescents and young adults’ relationships, life values, personal meaning, or coping strategies^75^. Moreover, the findings of the study may help to develop personalized preventive or therapeutic interventions, enhancing youths’ ability to cope with daily stress and increase their resilience through the reciprocal link between spirituality and positive or negative emotions. The study’s result may also raise awareness among the general population about the role of spirituality and self-regulation in mental health, and how they can be used to improve well-being. The new and unexpected findings of this study may provide valuable insights for conducting larger-scale research with more a priori hypotheses in contrast to the more explorative nature of the current study.

## Data Availability

No applicable

## Declarations

### Contributors

SU, JG & KJP designed the project. SR prepared the planned analyses. JG obtained the grant. SU, CN & JG drafted the present manuscript. All authors critically revised the manuscript and approved the final version of the manuscript.

### Patient and public involvement

Youth contributed to the design and protocol development, particularly for the ambulatory assessments. Further details are provided in the Methods section.

### Patient consent for publication

Not applicable

## References

1. Cangas AJ, Navarro N, Parra J, et al. Stigma-Stop: A serious game against the stigma toward mental health in educational settings. Frontiers in psychology. 2017;8:1385.

2. Goldbeck L, Schmitz TG, Besier T, Herschbach P, Henrich G. Life satisfaction decreases during adolescence. Qual Life Res. Aug 2007;16(6):969–79. doi:10.1007/s11136-007-9205-5

3. Maizes V, Rakel D, Niemiec C. Integrative medicine and patient-centered care. Explore (NY). Sep-Oct 2009;5(5):277-89. doi:10.1016/j.explore.2009.06.008

4. Sulmasy DP. A biopsychosocial-spiritual model for the care of patients at the end of life. Gerontologist. 2002;3:24–33. doi:10.1093/geront/42.suppl_3.24. PMID: 12415130

5. Hefti R. Integrating Religion and Spirituality into Mental Health Care, Psychiatry and Psychotherapy. Religions. 2011;2:611–627. doi:10.3390/rel2040611

6. Cabitza M, Riedo G, Bosisio F, et al. *« Donner la voix à la population » Enquête auprès de la population romande concernant la santé intégrative* Initiative Santé intégrative et société 2022.

7. Roth M, Müller F. *Pratiques et croyances religieuses et spirituelles en Suisse: Premiers résultats de l’Enquête sur la langue, la religion et la culture* 2019. Département Fédéral de l’Intérieur: Office Fédéral de la Statistique 2020.

8. statistique Ofdl. Religiosité et spiritualité en Suisse. 2025;doi:10.71668/nrv7-ea03

9. Pargament KI, A. M, Exline JJ, W. JJ, P. SE. Envisioning an integrative paradigm for the psychology of religion and spirituality. In: Pargament KI, Exline JJ, Jones WJ, eds. APA handbook of psychology, religion, and spirituality. American Psychological Association; 2013:3–19:chap Context, theory, and research.

10. Delaney C. The Spirituality Scale: development and psychometric testing of a holistic instrument to assess the human spiritual dimension. J Holist Nurs. Jun 2005;23(2):145–67; discussion 168-71. doi:10.1177/0898010105276180

11. Koenig HG, Zaben FA, Khalifa DA. Religion, spirituality and mental health in the West and the Middle East. Asian J Psychiatr. Jun 2012;5(2):180–2. doi:10.1016/j.ajp.2012.04.004

12. Aldwin CM, Park CL, Jeong Y-J, Nath R. Differing pathways between religiousness, spirituality, and health: A self-regulation perspective. Psychology of Religion and Spirituality. 2014;6:9–21. doi:10.1037/a0034416

13. Bozek A, Nowak PF, Blukacz M. The Relationship Between Spirituality, Health-Related Behavior, and Psychological Well-Being. Front Psychol. 2020;11:1997. doi:10.3389/fpsyg.2020.01997

14. Fredrickson BL. How does religion benefit health and wellbeing?: Are positive emotions active ingredients? Psychological Inquiry. 2002;13:209–213.

15. Pargament KI. Spiritually integrated psychotherapy: Understanding and addressing the sacred. Guilford Press; 2007.

16. Hardy SA, Nelson JM, Moore JP, King PE. Processes of Religious and Spiritual Influence in Adolescence: A Systematic Review of 30 Years of Research. J Res Adolesc. Jun 2019 2019;29(2):254–275. doi:10.1111/jora.12486

17. Ironson G, Stuetzle R, Fletcher MA. An increase in religiousness/spirituality occurs after HIV diagnosis and predicts slower disease progression over 4 years in people with HIV. J Gen Intern Med. Dec 2006;21 Suppl 5(Suppl 5):S62–8. doi:10.1111/j.1525-1497.2006.00648.x

18. Oman D, Thoresen CE. Do religion and spirituality influence health?. In: Paloutzian RF, Park CL, eds. Handbook of the psychology of religion and spirituality. Guilford Press; 2005:435–459.

19. Garssen B, Visser A, Pool G. Does Spirituality or Religion Positively Affect Mental Health? Meta-analysis of Longitudinal Studies. The International Journal for the Psychology of Religion. 2021;31:4–20. doi:10.1080/10508619.2020.1729570

20. James AG, Miller B. Revisiting Mahoney’s ‘My body is a temple’study: Spirituality as a mediator of the religion–health interaction among adolescents. International Journal of Children’s Spirituality. 2017;22:134–153. doi:10.1080/1364436X.2017.1301888

21. Wong YJ, Rew L, Slaikeu KD. A systematic review of recent research on adolescent religiosity/spirituality and mental health. Issues Ment Health Nurs. Feb-Mar 2006;27(2):161–83. doi:10.1080/01612840500436941

22. Kao LE, Peteet JR, Cook CC. Spirituality and mental health. Journal for the Study of Spirituality. 2020;10:42–54. doi:10.1080/20440243.2020.1726048

23. Kim S, Miles-Mason E, Kim CY, Esquivel GB. Religiosity/spirituality and life satisfaction in Korean American adolescents. Psychology of Religion and Spirituality. 2013;5:33. doi:10.1037/a0030628

24. Marques SC, Lopez SJ, Mitchell J. The role of hope, spirituality and religious practice in adolescents’ life satisfaction: Longitudinal findings. Journal of Happiness Studies. 2013;14:251–261. doi:10.1007/s10902-012-9329-3

25. Cotton S, Zebracki K, Rosenthal SL, Tsevat J, Drotar D. Religion/spirituality and adolescent health outcomes: a review. J Adolesc Health. Apr 2006;38(4):472–80. doi:10.1016/j.jadohealth.2005.10.005

26. Brandt P-Y. *Introduction à la psychologie de la religion*. Labor et Fides; 2023.

27. Nigg JT. Annual Research Review: On the relations among self-regulation, self-control, executive functioning, effortful control, cognitive control, impulsivity, risk-taking, and inhibition for developmental psychopathology. Journal of Child Psychology and Psychiatry. 2017;58(4):361–383. doi:10.1111/jcpp.12675

28. Shroff DM, Breaux R, von Suchodoletz A. Understanding the association between spirituality and mental health outcomes in adolescents in two non-Western countries: Exploring self-control as a potential mediator. Dev Psychopathol. Dec 23 2021:1–10. doi:10.1017/S0954579421001334

29. Barton KS, Tate T, Lau N, Taliesin KB, Waldman ED, Rosenberg AR. "I’m Not a Spiritual Person." How Hope Might Facilitate Conversations About Spirituality Among Teens and Young Adults With Cancer. J Pain Symptom Manag. Jun 2018;55(6):1599–1608. doi:10.1016/j.jpainsymman.2018.02.001

30. Farahani AS, Rassouli M, Yaghmaie F, Majd HA. Hope, the Foundation of Spiritual Health in Adolescents: A Qualitative Study. Iran Red Crescent Me. Dec 2016;18(12)doi:ARTN e29328 10.5812/ircmj.29328

31. Pour N, Mahmoodi-Shan G, Ebadi A, Behnampour N. Spiritual self-care in adolescents: a qualitative study. International Journal of Adolescent Medicine and Health. 2020;34:49–57. doi:10.1515/ijamh-2019-0248

32. Toruner EK, Altay N, Ceylan C, Arpaci T, Sari C. Meaning and Affecting Factors of Spirituality in Adolescents. Journal of Holistic Nursing. Dec 2020;38(4):362–372. doi:Artn 0898010120920501 10.1177/0898010120920501

33. Baumann M, Khaliefi RC. Muslim and Buddhist Youths in Switzerland: Individualising Religion and Striving for Recognition? Soc Incl. 2020;8(3):273–285. doi:10.17645/si.v8i3.3071

34. Demmrich S, Huber S. Multidimensionality of Spirituality: A Qualitative Study among Secular Individuals. Religions. Nov 2019;10(11)doi:ARTN 613 10.3390/rel10110613

35. Shiffman S, Stone AA, Hufford MR. Ecological momentary assessment. Annu Rev Clin Psycho. 2008;4:1–32. doi:10.1146/annurev.clinpsy.3.022806.091415

36. Balkaya-Ince M, Tahseen M, Umarji O, Schnitker SA. Does ramadan serve as a naturalistic intervention to promote muslim american adolescents’ daily virtues? Evidence from a three wave experience sampling study. The Journal of Positive Psychology. 2023;doi:10.1080/17439760.2023.2169631

37. Brelsford GM, Underwood LG, Wright BRE. Love in the midst of stressors: Exploring the role of daily spiritual experiences. In: Hood RW, Cheruvallil-Contractor S, eds. Research in the social scientific study of religion. BRILL; 2019:25-43.

38. Kent BV. Accelerated and micro-longitudinal approaches to understanding depressive symptoms and human flourishing. Ph.D. Baylor University; 2018.

39. Kent BV, Henderson MW, Bradshaw M, Ellison CG, Wright BRE. Do Daily Spiritual Experiences Moderate the Effect of Stressors on Psychological Well-being? A Smartphone-based Experience Sampling Study of Depressive Symptoms and Flourishing. The International Journal for the Psychology of Religion. 2021;31(2):57–78. doi:10.1080/10508619.2020.1777766

40. Kucinskas J, Wright BRE, Ray DM, Ortberg J. States of Spiritual Awareness by Time, Activity, and Social Interaction. Journal for the Scientific Study of Religion. 2017;56(2):418–437.

41. Kucinskas J, Wright BRE, Riepl S. The interplay between meaning and sacred awareness in everyday life: Evidence from a daily smartphone study. The International Journal for the Psychology of Religion. 2018;28(2):71–88. doi:10.1080/10508619.2017.1419050

42. Olson R, Knepple Carney A, Hicks Patrick J. Associations between gratitude and spirituality: An experience sampling approach. Psychology of Religion and Spirituality. 2019;11(4):449–452. doi:10.1037/rel0000164

43. van den Brink B, Jongkind M, Wijzenbroek W, et al. The Experience Sampling Method: A New Way of Assessing Variability of the Emotional Dimensions of Religiosity and Spirituality in a Dutch Psychiatric Population. J Relig Health. Oct 2023;62(5):3687–3701. doi:10.1007/s10943-023-01857-w

44. Wilt JA, Exline JJ, Pargament KI. Daily measures of religious/spiritual struggles: Relations to depression, anxiety, satisfaction with life, and meaning. Psychology of Religion and Spirituality. 2021;doi:10.1037/rel0000399

45. Grossoehme DH, Szczesniak RD, Mrug S, Dimitriou SM, Marshall A, McPhail GL. Adolescents’ spirituality and cystic fibrosis airway clearance treatment adherence: Examining mediators. Journal of Pediatric Psychology. 2016;41(9):1022–1032. doi:10.1093/jpepsy/jsw024

46. Kopp AT, Chini BA, Dimitriou SM, Grossoehme DH. Body Sanctification and Sleep in Adolescents with Cystic Fibrosis: A Pilot Study. J Relig Health. Oct 2017;56(5):1837–1845. doi:10.1007/s10943-017-0415-z

47. Iannello NM, Inguglia C, Silletti F, et al. How Do Religiosity and Spirituality Associate with Health-Related Outcomes of Adolescents with Chronic Illnesses? A Scoping Review. Int J Env Res Pub He. Oct 2022;19(20)doi:ARTN 13172 10.3390/ijerph192013172

48. Lun VMC, Bond MH. Examining the Relation of Religion and Spirituality to Subjective Well-Being Across National Cultures. Psychology of Religion and Spirituality. Nov 2013;5(4):304–315. doi:10.1037/a0033641

49. Traphagan J. Multidimensional measurement of religiousness/spirituality for use in health research in cross-cultural perspective. Res Aging. Jul 2005;27(4):387–419. doi:10.1177/0164027505275193

50. Harris PA, Taylor R, Minor BL, et al. The REDCap consortium: Building an international community of software platform partners. J Biomed Inform. Jul 2019;95:103208. doi:10.1016/j.jbi.2019.103208

51. Harris PA, Taylor R, Thielke R, Payne J, Gonzalez N, Conde JG. Research Electronic Data Capture (REDCap) - a metadata-driven methodology and workflow process for providing translational research informatics support. J Biomed Inform. Apr 2009;42(2):377–81. doi:10.1016/j.jbi.2008.08.010

52. Preventing Adverse Childhood Experiences: Leveraging the Best Available Evidence (2019).

53. Thakur N, Hessler D, Koita K, et al. Pediatrics adverse childhood experiences and related life events screener (PEARLS) and health in a safety-net practice. Child Abuse Negl. Oct 2020;108:104685. doi:10.1016/j.chiabu.2020.104685

54. Labelle R, Breton JJ, Berthiaume C, et al. Psychometric properties of three measures of protective factors for depression and suicidal behaviour among adolescents. Can J Psychiatry. Feb 2015;60(2 Suppl 1):S16–26.

55. Mirkovic B, Belloncle V, Pellerin H, Guile JM, Gerardin P. Gender Differences Related to Spirituality, Coping Skills and Risk Factors of Suicide Attempt: A Cross-Sectional Study of French Adolescent Inpatients. Front Psychiatry. 2021;12:537383. doi:10.3389/fpsyt.2021.537383

56. Braghetta CC, Gorenstein C, Wang YP, et al. Development of an Instrument to Assess Spirituality: Reliability and Validation of the Attitudes Related to Spirituality Scale (ARES). Front Psychol. 2021;12:764132. doi:10.3389/fpsyg.2021.764132

57. Steger MF, Frazier P, Oishi S, Kaler M. The Meaning in Life Questionnaire: Assessing the presence of and search for meaning in life. Journal of Counseling Psychology. 2006;53(80-93)

58. d’Acremont M, Van der Linden M. How is impulsivity related to depression in adolescence? Evidence from a French validation of the cognitive emotion regulation questionnaire. Validation Studies. J Adolesc. Apr 2007;30(2):271–82. doi:10.1016/j.adolescence.2006.02.007

59. Brevers D, Foucart J, Verbanck P, Turel O. Examination of the validity and reliability of the French version of the Brief Self-Control Scale. Can J Behav Sci. Oct 2017;49(4):243–250. doi:10.1037/cbs0000086

60. d’Acremont M, Van der Linden M. Adolescent Impulsivity: Findings from a Community Sample. Journal of Youth and Adolescence. 2005;34:427.

61. Billieux J, Rochat L, Ceschi G, et al. Validation of a short French version of the UPPS-P Impulsive Behavior Scale. Compr Psychiatry. Jul 2012;53(5):609–15. doi:10.1016/j.comppsych.2011.09.001

62. Dumont M, Schwarzer R, Jerusalem M. French adaptation of the general self-efficacy scale http://userpage.fu-berlin.de/~health/french.htm

63. Vermeersch S, Fombonne E. Le Child Behavior Checklist: Résultats préliminaires de la standardisation de la version française. *Neuropsychiatrie de l’* Enfance et de l’ Adolescence. 1997;45:615–620.

64. Vreugdenhil C, van den Brink W, Ferdinand R, Wouters L, Doreleijers T. The ability of YSR scales to predict DSM/DISC-C psychiatric disorders among incarcerated male adolescents. Eur Child Adolesc Psychiatry. Mar 2006;15(2):88–96. doi:10.1007/s00787-006-0497-8

65. Lukat J, Margraf J, Lutz R, van der Veld WM, Becker ES. Psychometric properties of the Positive Mental Health Scale (PMH-scale). BMC Psychol. Feb 10 2016;4:8. doi:10.1186/s40359-016-0111-x

66. Herdman M, Rajmil L, Ravens-Sieberer U, Bullinger M, Power M, Alonso J. Expert consensus in the development of a European health-related quality of life measure for children and adolescents: a Delphi study. Acta Paediatrica. 2002;91(12):1385–1390.

67. Rajmil L, Herdman M, de Sanmamed M-J, et al. Generic health-related quality of life instruments in children and adolescents: a qualitative analysis of content. Journal of adolescent Health. 2004;34(1):37–45.

68. Robitail S, Simeoni M-C, Erhart M, et al. Validation of the European proxy KIDSCREEN-52 pilot test health-related quality of life questionnaire: first results. Journal of Adolescent Health. 2006;39(4):596. e1-596. e10.

69. Lamers F, Swendsen J, Cui L, et al. Mood reactivity and affective dynamics in mood and anxiety disorders. J Abnorm Psychol. Oct 2018;127(7):659–669. doi:10.1037/abn0000378

70. Drexl K, Urben S, von Plessen KJ, Glaus J. Using mobile assessments to characterize mental and physical health behaviors in youth: Protocol for a pilot observational intensive longitudinal study. JMIR Res Protoc. 2024;14:e70990.

71. Plessen KJ, Constanty L, Ranjbar S, et al. The role of self-regulatory control processes in understanding aggressive ideations and behaviors: An experience sampling method study. Front Psychiatry. 2022;13:1058814. doi:10.3389/fpsyt.2022.1058814

72. Braun V, Clarke V. Using thematic analysis in psychology. Qualitative Research in Psychology. 2006/01/01 2006;3(2):77–101. doi:10.1191/1478088706qp063oa

73. R Core Team. R: A language and environment for statistical computing. R Foundation for Statistical Computing; 2021.

74. SAS Institute Inc. Logiciel SAS (Version 9.4). SAS Institute Inc. 2023;

75. Smith BW, Ortiz JA, Wiggins KT, Bernard JF, Dalen J. Spirituality, resilience, and positive emotions. In: Miller LJ, ed. The Oxford Handbook of Psychology and Spirituality. Oxford University Press; 2012:437–454.

76. Tasse MJ, Balboni G, Navas P, et al. Developing behavioural indicators for intellectual functioning and adaptive behaviour for ICD-11 disorders of intellectual development. J Intellect Disabil Res. May 2019;63(5):386–407. doi:10.1111/jir.12582

77. WHO WHO. International classification of diseases for mortality and morbidity statistics: 11th revision. 2018.

